# The impact of introduction of the 10-valent pneumococcal conjugate vaccine (PCV10) on pneumococcal carriage in Nigeria

**DOI:** 10.1101/2022.03.11.22271682

**Authors:** Aishatu L Adamu, J. Ojal, Isa A. Abubakar, Kofo Odeyemi, Musa M. Bello, Christy A.N. Okoromah, Boniface Karia, Angela Karani, Donald. Akech, Victor Inem, J. Anthony G Scott, Ifedayo M.O. Adetifa

## Abstract

**Background:** Pneumococcal Conjugate Vaccines (PCVs) reduce the burden of pneumococcal disease by reducing nasopharyngeal acquisition and transmission of serotypes in the vaccines (vaccine-serotypes). Following introduction of the 10-valent PCV (PCV10) in Nigeria, we assessed the population-level impact of PCV introduction on pneumococcal carriage.

**Methods:** We conducted annual cross-sectional carriage and vaccination coverage surveys between 2016 and 2020 in rural and urban sites in Nigeria. We recruited a random sample of residents and used WHO-recommended laboratory methods to identify pneumococcal serotypes. We modelled prevalence ratios for the change in annual carriage prevalence using log-binomial regression, and explored the association between vaccination coverage in children <5 years and changes in the population-level vaccine-serotype carriage.

**Findings:** We enrolled 4,684 and 3,653 participants for the carriage surveys and 2,135 and 1,106 children for the coverage surveys in the rural and urban sites, respectively. Carriage prevalence of vaccine-serotypes declined steadily with increasing levels of PCV10 coverage among children aged <5 years. From 2016 to 2020, coverage with three doses of PCV10 increased from 7% to 59% and from 15% to 81% in the rural and urban sites, respectively. Prevalence ratios for the annual change in vaccine-serotype carriage in participants aged <5 years and ≥5 years were 0.84 (95% CI:0.79-0.89) and 0.86 (95%CI:0.80-0.89) in the rural, and 0.69 (95% CI:0.62-0.77), and 0.83 (95% CI:0.74-0.93) in the urban sites.

**Conclusions:** We found significant reduction in vaccine-serotype carriage at a population level following a steady increase in PCV10 coverage, indicating direct and indirect vaccine effects.

## Introduction

The widespread introduction of pneumococcal conjugate vaccines (PCVs) has resulted in a substantial decline in the burden of pneumonia and invasive pneumococcal disease (IPD) in children.[1] PCV use has also substantially reduced hospital visits from pneumococcal otitis media and antibiotic use and has the potential to reduce antimicrobial resistance.[2–4] Nonetheless, *Streptococcus pneumoniae* (the pneumococcus) remains a leading cause of pneumonia and invasive diseases (meningitis, bacteraemic pneumonia and bacteraemia).[1] In 2015, pneumococcal disease still caused approximately 300,000 deaths among children aged 1-59 months globally, and over 50% of these deaths occurred in the African sub-region. Nigeria alone accounted for nearly 50,000 of these deaths.[1]

Nasopharyngeal carriage precedes pneumococcal disease.[5] Young children are the main reservoirs for carriage and accordingly have the highest burden of pneumococcal disease.[5] PCVs directly protect vaccinated children against acquisition of carriage and disease from the vaccine-type (VT) and indirectly protect unvaccinated persons by preventing carriage and transmission of these serotypes.[6,7] This indirect impact accounts for a substantial fraction of the overall PCV impact.[6] The direct and indirect effectiveness of PCV10 against pneumococcal carriage and disease has been demonstrated in various settings.[2,7] The PCV-induced decline of VT-serotype carriage and in IPD burden is associated with replacement by non-vaccine serotypes (NVTs) in carriage and, to a varying extent, with serotype replacement disease (SRD) across a variety of settings.[7–9]

Between 2014 and 2016, Nigeria introduced the 10-valent PCV (PCV10) for infants without a catch-up campaign. Prior to PCV use, the serotypes included in PCV10 accounted for approximately 72% of IPD among children less than five years in Africa.[10] The net benefit of PCV at the population level depends on serotype distribution in carriage and disease, transmission intensity or force of infection, vaccination coverage and serotype replacement.[5,11,12] A substantial component of PCV’s impact relies on protection against pneumococcal carriage.[5] Carriage studies conducted at the time of PCV10 introduction in Nigeria among children aged <5 years showed overall carriage of 71% and 92% and VT carriage of 46% and 50%, in urban and rural settings, respectively.[13] Carriage prevalence was also substantial in persons aged ≥5 years at >40% and >35%, respectively in these settings, and VT carriage accounted for nearly half of all observed carriage.[13]

To evaluate the impact of introducing PCV10 on pneumococcal carriage and serotype distribution, we conducted annual carriage and vaccination coverage surveys in two sites in Nigeria. We assessed and described changes in overall, VT, and NVT carriage in children aged <5 years and persons aged ≥5 years following PCV10 introduction, and explored the relation between these and vaccination coverage.

## Methods

### Study design and participants

We conducted annual cross-sectional carriage and vaccine coverage surveys in Kumbotso, Kano State (rural) and Pakoto, Ogun State (urban). PCV10 was introduced in Kumbotso in July 2016 and in Pakoto in October 2016 with a schedule of three primary doses (3p+0) at ages 6, 10 and 14 weeks with no catch-up for older children. The catchment areas for the carriage and vaccine coverage surveys were those within 10km of the Kumbotso and Pakoto Comprehensive Primary Health Care Centres in the rural and urban sites, respectively. Baseline carriage surveys were conducted in December 2016 (rural) and February 2017 (urban), four to five months after PCV10 was introduced.[13]

Surveys were repeated annually using the same design in November/December for four years (2017-2020) in the rural site and in February/March for three years (2018-2020) in the urban site (Fig S1). The target population for the carriage and vaccine coverage surveys were all residents of the respective communities living within the catchment areas. We used a two-stage sampling design for each survey between 2017 and 2020 to select participants at random. In the first stage, we conducted a census of all households within 10km of the two health centres and assigned each household a unique identification number. We then selected households using simple random sampling.

In the second stage of the carriage surveys, we randomly chose one participant per selected household systematically drawn from increasing age-strata at each new household. We recruited participants in ten age groups (<1, 1-2, 3-4, 5-9, 10-14, 15-19, 20-39, 40-49, 50-59, and ≥ 60 years), starting with the lowest and moving to the highest age group until we had recruited one participant per age group and then we restarted the process. If there was no participant in a particular age group in the household, we selected the next age group in sequence and then looked for any missed age groups in subsequent households.

A total of 1,000 participants per carriage survey, with a target of 100 per age group, was expected to give at least 90% power to detect a 50% decline in VT carriage from a baseline of 22-26%.[13]

We conducted three PCV10 coverage surveys in the rural site and two in the urban site between 2018 and 2020 (figure S1). The target population for PCV10 coverage surveys were resident children <5 years old who were age-eligible to have received three doses of PCV10 at the date of the first post-PCV survey. For the PCV10 coverage surveys, households were independently selected at random from the census described earlier. In the second stage, all eligible children per selected household were recruited.

Assuming at least two eligible children per household and 80% response or participation, a sample size of 639 children per site per survey was sufficient to estimate coverage of the third dose of PCV of 50% with a 5% precision. Targeting a vaccination coverage of 50% allowed estimation of the largest possible sample size.

### Procedure

Sociodemographic and clinical information was obtained from participants of the carriage surveys using an interviewer-administered questionnaire. Nasopharyngeal swabbing, transport, storage and culture were done according to WHO recommended standards.[14] We collected one swab specimen per participant from the posterior wall of the nasopharynx using nylon-tipped flexible flocked swabs. We immediately inserted the swab into a tube containing skimmed milk-tryptone-glucose-glycerin (STGG) by cutting the wire portion of the swab just above the top of the tube. We placed STGG tubes containing the swabs on ice packs in a cold box, and they were transported within 8 hours of collection for temporary storage at temperatures between -80°C and -55°C. We later shipped swabs on dry ice to the KEMRI-Wellcome Trust Research Programme (KWTRP), Kilifi, Kenya, and stored them at -80°C until they were processed. We identified pneumococci by culture using α-haemolysis and optochin sensitivity testing. We identified serotypes using latex agglutination and Quellung Reaction. For isolates with inconclusive Quellung reaction, polymerase chain reaction (PCR) targeting the genes encoding autolysin (lytA) and multiplex PCR were used to confirm the isolate as pneumococcus and for serotyping.

For the PCV10 coverage survey, we visited selected households to interview caregivers. We then obtained the vaccination status of each child, including doses and dates received from the vaccination cards or caregiver recall and recorded in the questionnaire.

### Statistical analysis

#### Carriage surveys

We calculated the age-stratified and population-based carriage prevalence for each survey year for all pneumococci, VT, and NVT. VT carriage was defined as carriage of pneumococcal serotypes in PCV10 (1, 4, 5, 6B, 7F, 9V, 14, 18C, 19F and 23F), while NVT carriage was carriage of any non-PCV10 serotype, including non-typeable isolates.

We used log-binomial regression with robust standard errors to model changes in carriage prevalence over time as prevalence ratios (PRs) and used Poisson regression when the models failed to converge. We estimated PRs with 95% confidence intervals (CIs) over time, in years, from PCV10 introduction to date of swab collection. We adjusted PRs for exposure variables that were separately associated with carriage and survey year at p <0.1 (living with children aged <5 years and history of upper respiratory tract infection symptoms). We also adjusted for the stratified sampling method by standardising PRs in the sampling age strata to the age distribution of Kumbotso LGA (for rural) and Ifo and Ado-Ota LGAs (for urban) obtained from population models of Nigerian census data.[15] We calculated overall PRs, and age-specific PRs for children aged <5 years and persons aged ≥5 years. To summarise the total impact of the vaccine programme, we compared adjusted carriage prevalence in the final survey to carriage prevalence in the first survey.

#### Vaccination coverage surveys

We assessed PCV10 coverage in each survey year (2018-2020) as the proportion of children who received two doses of PCV10 irrespective of timing and age of receipt. Because we did not conduct PCV10 coverage surveys in the early period (2016-2017), we used a birth cohort analysis approach to estimate the PCV10 coverage of children aged <5years for these years from the data collected in 2018-20. In addition, we adjusted for clustering at the household level to account for non-independence of observations for children sampled within the same household.

We compared the annual coverage with three doses of PCV10 in children <5 years to VT carriage in children aged <5 years and persons aged ≥5 years.

We did all analyses with Stata® version 15.1(College Station, Texas, United States) and conducted the analysis separately for each site.

## Results

We conducted five annual carriage surveys in the rural and four in the urban sites (S1 Fig) and recruited 4,684 and 3,653 participants, respectively. In rural and urban sites, the proportion of potential participants who consented to be in the swabbed varied from 60-98% and 63-99%, respectively, across the sampling age groups and surveys (S1 Table and S2 Fig).

Participants in the rural site resided in larger households and more commonly reported living with ≥2 children aged <5 years, using solid fuel for cooking, and having a cough or runny nose in the preceding two weeks compared to their counterparts in the urban site (Table 1).

**Table 1:** Background characteristics of study participants.

|  | Survey 1 | Survey 2 | Survey 3 | Survey 4 | Survey 5 |
| --- | --- | --- | --- | --- | --- |
| **Rural site** |  |  |  |  |  |
| Total sample | 878 | 879 | 999 | 973 | 954 |
| **Age (years)** |  |  |  |  |  |
| <5 | 296 (34%) | 264 (30%) | 304 (30%) | 365 (37%) | 333 (35%) |
| 5-9 | 191 (22%) | 116 (13%) | 102 (10%) | 124 (13%) | 120 (13%) |
| 10-19 | 119 (13%) | 216 (25%) | 216 (22%) | 202 (21%) | 234 (24%) |
| 20-39 | 154 (17%) | 104 (12%) | 125 (12%) | 101 (10%) | 122 (13%) |
| ≥40 | 112 (13%) | 178 (20%) | 252 (25%) | 184 (19%) | 144 (15%) |
| **Clinical history** |  |  |  |  |  |
| Runny nose | 714 (81%) | 681 (77%) | 900 (90%) | 843 (87%) | 727 (76%) |
| Cough | 450 (51%) | 551 (63%) | 687 (69%) | 558 (57%) | 487 (51%) |
| Antibiotic use | 65 (7%) | 431 (49%) | 510 (51%) | 233 (24%) | 202 (21%) |
| **Household composition** |  |  |  |  |  |
| Living with ≥2 under-fives | 645 (73%) | 469 (53%) | 555 (56%) | 619 (64%) | 748 (78%) |
| Sharing bed with ≥2 persons | 729 (83%) | 704 (80%) | 882 (88%) | 795 (82%) | 857 (90%) |
| **Household cooking fuel** |  |  |  |  |  |
| Solid fuel | 833 (95%) | 795 (90%) | 959 (96%) | 892 (92%) | 850 (89%) |
| **Household size** |  |  |  |  |  |
| All persons, median (IQR) | 9 (7-13) | 6 (3-10) | 6 (4-9) | 8 (6-10) | 9 (7-12) |
| **Urban site** |  |  |  |  |  |
| Total sample | 924 | 943 | 932 | 854 | N/A |
| **Age (years)** |  |  |  |  |  |
| <5 | 335 (36%) | 244 (26%) | 243 (26%) | 185 (22%) | N/A |
| 5-9 | 161 (17%) | 119 (13%) | 117 (12%) | 116 (14%) | N/A |
| 10-19 | 95 (10%) | 209 (22%) | 178 (19%) | 193 (23%) | N/A |
| 20-39 | 130 (14%) | 91 (10%) | 106 (11%) | 104 (12%) | N/A |
| ≥40 | 198 (21%) | 278 (29%) | 288 (31%) | 243 (30%) | N/A |
| **Clinical history** |  |  |  |  |  |
| Runny nose | 238 (26%) | 163 (17%) | 106 (11%) | 51 (6%) | N/A |
| Cough | 216 (23%) | 122 (13%) | 75 (8%) | 32 (4%) | N/A |
| Antibiotic use | 145 (16%) | 76 (8%) | 39 (4%) | 10 (1%) | N/A |
| **Household composition** |  |  |  |  |  |
| Living with ≥2 under-fives | 95 (10%) | 81 (9%) | 69 (7%) | 53 (6%) | N/A |
| Sharing bed with ≥2 persons | 185 (20%) | 212 (23%) | 121 (13%) | 121 (14%) | N/A |
| **Household cooking fuel** |  |  |  |  |  |
| Solid fuel | 58 (6%) | 38 (4%) | 35 (4%) | 11 (1%) | N/A |
| **Household size** |  |  |  |  |  |
| All persons, median (IQR) | 4 (3-5) | 5 (4-6) | 5 (4-6) | 5 (4-6) | N/A |

The carriage prevalence across the sampled age groups is shown in S3 Fig. Overall carriage in the rural site was consistently high across all ages in all surveys. Table 2 shows the crude and age-standardised carriage prevalence stratified by survey. Carriage was higher in the children aged <5 years at both locations. For both age categories, the overall and NVT carriage prevalence were higher in the rural site. However, in both sites, and despite the initial rise in overall prevalence, there was a steady decline in VT carriage prevalence and an increase in NVT carriage prevalence in both age groups over the survey years (Chi squared test for trend, p <0·0001).

**Table 2:**
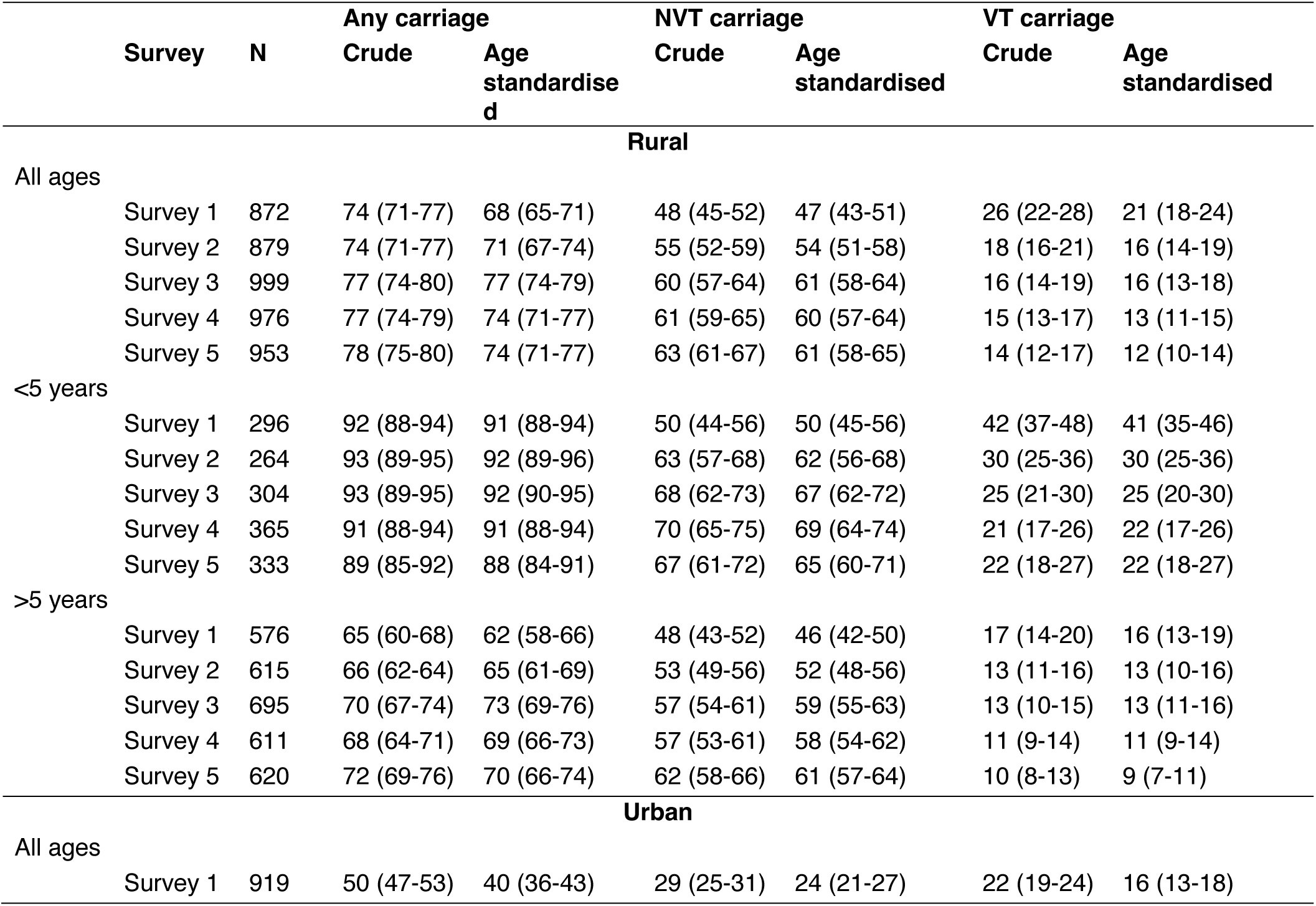
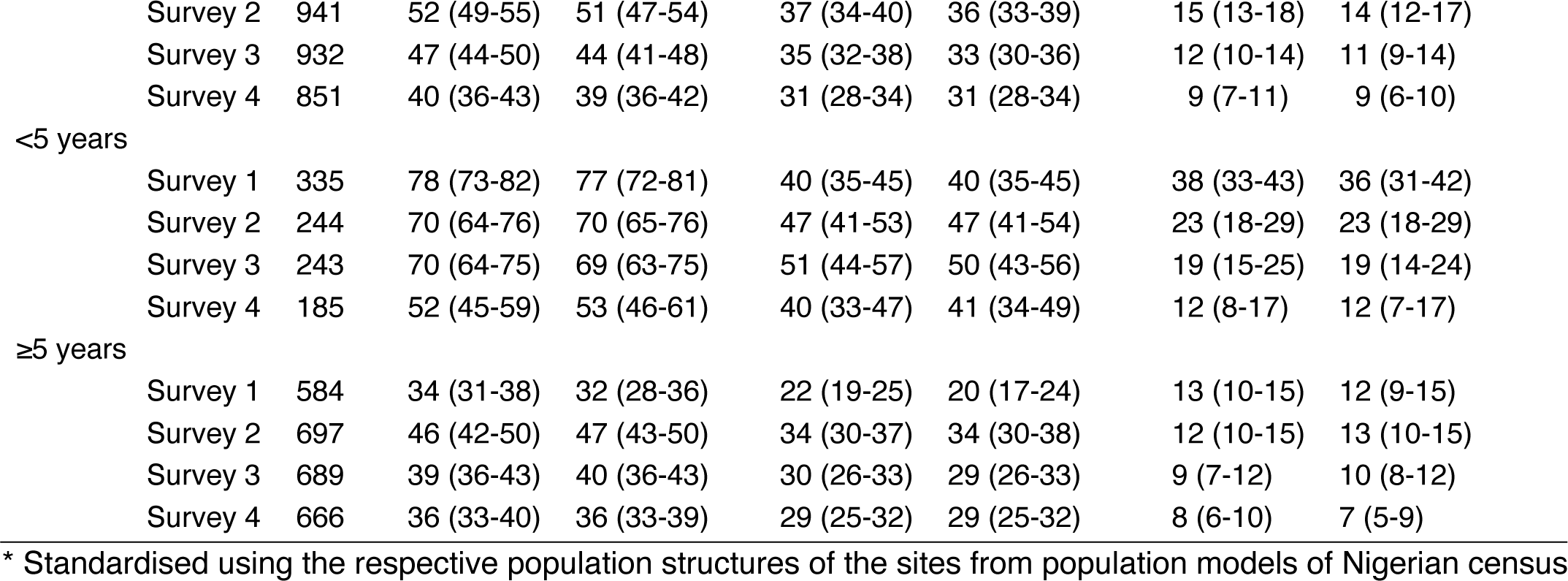
Crude and age-standardised* carriage prevalence of any, non-vaccine serotype (NVT), and vaccine serotype (VT) pneumococci stratified by site, age group, and survey.

Table 3 shows the crude and adjusted PRs for change in carriage prevalence per year and the overall change in carriage prevalence since PCV10 introduction. VT carriage declined by 16% per year (PR 0·84) in the rural and 31% (PR 069) in the urban sites in children aged <5 years. For participants, aged ≥5 years, the annual decline in VT carriage was 14% (PR 0·86) in the rural site and 17% (PR 0·83) in the urban site. When the final is compared to the first survey, the VT carriage prevalence declined in the two age groups by 48% and 47% in the rural site and by 69% and 40% in the urban site. respectively. Prevalence of NVT carriage also increased in both sites, except for children aged <5 years in the urban site where the 95% CIs were overlapping. Pneumococcal carriage prevalence overall, did not change in the rural site but declined by 9% per year (PR 0·91) in the urban site (Table 3) but this effect was restricted to children aged <5 years.

**Table 3:** Prevalence ratios (PR), and 95% CI, showing changes in any, non-vaccine serotype (NVT), and vaccine serotype (VT) carriage stratified by age and site.

|  | Any carriage |  | NVT carriage |  | VT carriage |  |
| --- | --- | --- | --- | --- | --- | --- |
|  | Crude PR | Adjusted PR<sup>1</sup> | Crude PR | Adjusted PR<sup>1</sup> | Crude PR | Adjusted PR<sup>1</sup> |
| **PR for average change in carriage per year<sup>2</sup>** |  |  |  |  |  |  |
| Rural<sup>3</sup> |  |  |  |  |  |  |
| All ages | 1.01 (1.00-1.03) | 1.00 (0.99-1.02) | 1.06 (1.05-1.08) | 1.06 (1.04-1.08) | 0.86 (0.82-0.90) | 0.85 (0.81-0.89) |
| <5 years | 0.99 (0.96-1.03) | 0.99 (0.88-1.00) | 1.07 (1.04-1.10) | 1.07 (1.04-1.10) | 0.83 (0.79-0.88) | 0.84 (0.79-0.89) |
| ≥5 years | 1.03 (0.99-1.06) | 1.01 (0.99-1.03) | 1.06 (1.04-1.09) | 1.05 (1.03-1.08) | 0.88 (0.82-0.94) | 0.86 (0.80-0.92) |
| Urban<sup>4</sup> |  |  |  |  |  |  |
| All ages | 0.93 (0.90-0.96) | 0.91 (0.88-0.94) | 1.02 (0.98-1.06) | 1.02 (0.98-1.07) | 0.74 (0.69-0.80) | 0.69 (0.63-0.75) |
| <5 years | 0.90 (0.87-0.94) | 0.91 (0.88-0.94) | 1.03 (0.97-1.08) | 1.05 (0.98-1.12) | 0.69 (0.62-0.77) | 0.69 (0.62-0.77) |
| ≥5 years | 0.99 (0.95-1.03) | 1.00 (0.95-1.05) | 1.05 (0.99-1.11) | 1.06 (1.00-1.13) | 0.84 (0.76-0.94) | 0.83 (0.74-0.93) |
| **PR for overall change in carriage<sup>5</sup>** |  |  |  |  |  |  |
| Rural<sup>3</sup> |  |  |  |  |  |  |
| All ages | 1.06 (1.00-1.11) | 1.00 (0.95-1.05) | 1.32 (1.22-1.44) | 1.30 (1.19-1.42) | 0.55 (0.45-0.67) | 0.52 (0.43-0.64) |
| <5 years | 0.97 (0.82-1.14) | 0.97 (0.92-1.02) | 1.34 (1.17-1.54) | 1.34 (1.17-1.54) | 0.52 (0.41-0.67) | 0.52 (0.41-0.67) |
| ≥5 years | 1.12 (0.97-1.28) | 1.06 (0.97-1.14) | 1.31 (1.18-1.46) | 1.26 (1.12-1.40) | 0.58 (0.43-0.78) | 0.53 (0.39-0.72) |
| Urban<sup>4</sup> |  |  |  |  |  |  |
| All ages | 0.79 (0.71-0.88) | 0.72 (0.65-0.80) | 1.09 (0.95-1.26) | 1.03 (0.89-1.20) | 0.40 (0.31-0.51) | 0.34 (0.26-0.45) |
| <5 years | 0.67 (0.58-0.78) | 0.68 (0.58-0.79) | 1.01 (0.81-1.25) | 1.02 (0.82-1.28) | 0.32 (0.21-0.48) | 0.31 (0.20-0.48) |
| ≥5 years | 1.05 (0.91-1.22) | 1.07 (0.90-1.28) | 1.30 (1.07-1.58) | 1.36 (1.10-1.69) | 0.61 (0.44-0.86) | 0.60 (0.41-0.87) |

<sup>1</sup> adjusted for symptoms of upper respiratory tract infection in past two weeks, living with ≥2 under-fives, and age-standardised to age distribution of study sites; <sup>2</sup> prevalence ratios across all surveys from date of PCV10 introduction to date of swabbing coded as a single time variable to obtain estimate of prevalence ratio per year; <sup>3</sup> Five surveys (2016-2020); <sup>4</sup> Four surveys (2017-2020); <sup>5</sup> prevalence ratio comparing final survey in each site to the 1<sup>st</sup> survey.

Fig 1 shows the relationship between PCV10 coverage and annual VT carriage prevalence. VT carriage prevalence declined with increasing PCV10 coverage levels in both sites, particularly in children aged <5 years. The relationship between PCV10 coverage among children aged <5 years and VT carriage prevalence over time (Fig 2) shows the steepest decline in VT carriage occurred at coverage levels of <50% by the second year of PCV10 use. In 2020, there was a drop in PCV10 coverage in the rural site with an accompanying slight increase in VT carriage compared to values for the previous survey (2019).

The serotypes with the greatest reduction in carriage prevalence were 14, 19F, 23F and 6B. For children aged <5 years (S4 Fig and S2 Table), the serotypes with the highest age-standardised prevalence in the final surveys were 6A (11·4%), 19F (5·5%) and 19A (5·4%%), 11A (4·7%), 14 and 16F (4·4% each), and 23F (3·7%) in the rural site; and 19A (7·4%), 15B (4·6%), 6B (4·0%), 19F (3·9%), and 16F (3·7%) in the urban site. In persons aged ≥5 years NVTs dominated carriage in the final surveys (S5 Fig).

## Discussion

In this study, we determined the population-level impact of PCV10 introduction on pneumococcal carriage and related this to PCV10 coverage in children aged <5 years in the same populations. To our knowledge, this is the first study reporting the impact of PCV10 introduction on pneumococcal carriage in Nigeria. Our findings show significant changes in population pneumococcal carriage within four to five years of PCV10 introduction. In a rural and an urban site in Nigeria, with 52% and 64% cumulative vaccine coverage, respectively, VT carriage prevalence declined by 48% and 69% among children aged <5 years, and by 47% and 40% among persons aged ≥5 years. In contrast, NVT carriage increased significantly by 34% and 26% in the rural site for both age categories and by 36% but only in those aged ≥5 years in the urban site.

Although we observed a significant PCV10-induced decline in VT carriage in these Nigerian settings, the magnitude of the decline is lower than has been seen in settings that introduced PCV10 but achieved much higher vaccination coverage. In Kilifi, Kenya, VT carriage declined by 64% within the first year after PCV10 was introduced along with a catch-up campaign for all children <5 years along with the achievement of high vaccination coverage among infants (>80%) and children aged <5 years (67%) within a shorter period than observed in our study.[9,16] Similarly, in Brazil, three years after PCV10 was introduced with a catch-up for children 7-23 months, VT carriage declined by 91% in children 11-23 months that had achieved 95% coverage with 3-4 doses of PCV10. However, participant recruitment from immunisation clinics may have selected for children more likely to receive vaccination, thus overestimating vaccine impact.[17]

While the changes in carriage in the initial post-PCV period were modest, VT decline was substantial by the fourth year of PCV10 use in the urban site and similar to changes reported in Kilifi five years post-PCV10 introduction.[9] The relatively slower decline in our study is most likely due to the following: absence of catch-up vaccination to older children, the slow PCV10 uptake in both sites, and potential VT transmission from older children and adults, particularly in the context of high pre-PCV10 VT carriage in these groups and sub-optimal PCV10 coverage in the earlier study periods. Therefore, a longer period may be required before PCV10 produces further decline of VT carriage. Nonetheless, many African countries still report varying levels of residual VT carriage even after several years of high vaccine coverage.[9,18,19]

Although initiating the 3p+0 schedule early (6 weeks) and giving subsequent doses within short 4-week intervals aimed to quickly build up immunity early when children are most at risk, our findings still show substantial VT carriage beyond infancy. This residual VT carriage is more likely a consequence of high force of infection, contact patterns that enable effective transmission, or waning of vaccine-induced immunity.[18,20,21] A different PCV10 schedule including a booster dose, or mass PCV10 campaigns for older children might enhance indirect vaccine impact on VT carriage among infants, particularly given the substantial VT carriage in older children. For instance, some evidence suggests that a schedule with a booster dose after two primary doses (2p+1) enhances protection against VT carriage and longer intervals between doses elicit better immune response.[22] We note a dampening of vaccine impact and decline in PCV10 coverage in 2020 in the rural site, possibly because COVID19-related restrictions may have affected immunisation services. This rapid sensitivity to changes in PCV10 coverage is indicative of fragile herd immunity.

We observed overall significant carriage replacement with NVT in both locations in the final surveys. However, NVT replacement was limited to the population ≥5 years in the urban site. Although differences in the years of survey might explain some differences between the locations, when we considered each year of survey separately, NVT carriage (and pneumococcal carriage overall) and changes in NVT carriage were still significantly higher in the rural site. We attribute these differences to the higher carriage levels in all age groups, suggesting substantial transmission in older persons, and a larger pool of circulating serotypes in the rural compared to the urban site. The burden and distribution of NVT carriage in the population can influence serotype replacement if circulating NVTs have a competitive advantage for acquisition or evasion of clearance.[23] Varying levels of serotype replacement in carriage have been reported in other settings. After three years of PCV10 use in Fiji, NVT replacement was significant only in children <2 years but not for older children and caregivers.[24] Similarly, in Malawi, NVT replacement was only observed among fully vaccinated children 1-4 years after 2·5 years of PCV13 use.[25] In Kenya, six years post-PCV10 introduction, NVT replacement in carriage was significant in all age groups.[9]

Changes in carriage provide a valuable proxy for protection against disease because carriage is thought to be a necessary precursor for disease.[11,26] Furthermore, because both indirect and direct protection rely substantially on vaccine-induced protection against carriage, changes in carriage can provide useful insight into population-level vaccine impact on disease.[27] In Kenya, a 76% decline in VT carriage among children <5 years was associated with a 92% decline in VT IPD in this group and a smaller but significant decline in non-vaccine target age groups.[9] In Brazil, a 90% decline in VT carriage was associated with a VT-IPD decline of 83% in children <5 years and by ∼50% in older persons within five years of PCV10 introduction.[17,28] It would be reasonable to expect, therefore, that the decline in VT carriage we observed has translated to a proportional decline in IPD. The utility of simplified modelling approaches to predict PCV impact on IPD using carriage data has been demonstrated.[11,26]

Serotype replacement in carriage poses a potential threat to vaccine impact on IPD of all serotypes, particularly if replacement results from increased NVT acquisition of relatively invasive serotypes. However, because of the underlying high burden of NVT carriage in our study settings, replacement via unmasking from enhanced clearance or reduced carriage density in co-colonisation episodes is also a likely possibility.[23] In such circumstances, the risk of IPD may not be as high, particularly if these NVTs have an intrinsically low invasive capacity.[29] The high carriage burden in our study population may indicate a proportionate risk of IPD. However, it is worth noting that immunity levels needed to protect against invasive disease are lower than those required for protection against carriage.[5] Many African settings with longer PCV programmes have not reported significant SRD despite significant replacement in carriage.[8,9,26,30]

This study has some limitations. As we are evaluating an intervention in a longitudinal study, there may be some overlap between secular and vaccine-induced changes in carriage. To minimise confounding by seasonal changes in behaviour that may influence contact patterns and transmission risk, we conducted the surveys at similar times of year in each site, using the same field and laboratory methods. Of note, adjustment (for living with children aged <5 years, history of upper respiratory tract infection symptoms, and the age-stratified sampling) did not substantially alter the PRs. This finding suggests that our samples were largely similar across surveys with regards the proportion living with children aged <5 years, the proportion with URTI symptoms, and the proportion in each age stratum. We did not attempt to detect multiple serotype carriage in one swab; thus, we may have under-estimated carriage of some serotypes. The generalisability of our findings may be limited for Nigerian settings with different pneumococcal epidemiology. However, our study sites represent diverse settings sufficiently representative of most of Nigeria regarding risk factors for carriage, transmission and vaccine coverage.

In conclusion, PCV10 introduction in Nigeria, with accumulated vaccine uptake of three doses of PCV10 of 59-81% in the first five years, was associated with a significant decline in the prevalence of VT carriage among the vaccine-target and the non-target populations indicating substantial direct and indirect vaccine effects. These effects are likely to translate to a significant reduction in IPD burden. However, substantial residual carriage of vaccine serotypes possibly related to sub-optimal vaccine coverage and NVT replacement in carriage are potential threats to the PCV programme. Transmission dynamic modelling may provide better insight into the relationship between PCV coverage and population-level protection to define levels of coverage that would result in adequate herd protection and explore the potential of alternate schedules to reduce residual VT carriage further. Continuous monitoring of serotypes circulating in carriage can facilitate identification of emerging serotypes that may be important in SRD.

## Data Availability

All data produced in the present work are contained in the manuscript

## Authors’ contributions

ALA, IMOA, JAGS and JO contributed to study concept and design. ALA led the fieldwork with input from DA, AK, MMB, IAA, CANO, VI and KO. AK led the laboratory work. BA oversaw the curation and management of data. ALA performed all statistical analyses and interpreted the results with input from JO, IMOA and JAGS. ALA wrote the first draft. All authors contributed to critical revision of the manuscript for intellectual content, and had final responsibility for the decision to submit for publication.

## Data access

Data supporting findings are included in the appendix (S2 Table).

## Ethics

Written informed consent was obtained from participants/guardians. Ethics approval for study was granted by the Research Ethics Committees of Aminu Kano Teaching Hospital (NHREC/21/08/2008/AKTH/EC/2165), Kano State Ministry of Health (MOH/OFF/797/T.I/596), Lagos University Teaching Hospital (ADM/DCST/HREC/APP/10300); the Kenya Medical Research Institute’s Scientific and Ethical Review Unit (SERU 3350); and by the London School of Hygiene and Tropical Medicine Observational/Interventions Research Ethics Committee (Ref 11670).

