## Supplementary figures and images for "The impact of introduction of the 10-valent pneumococcal conjugate vaccine (PCV10) on pneumococcal carriage in Nigeria"

### S1 Fig

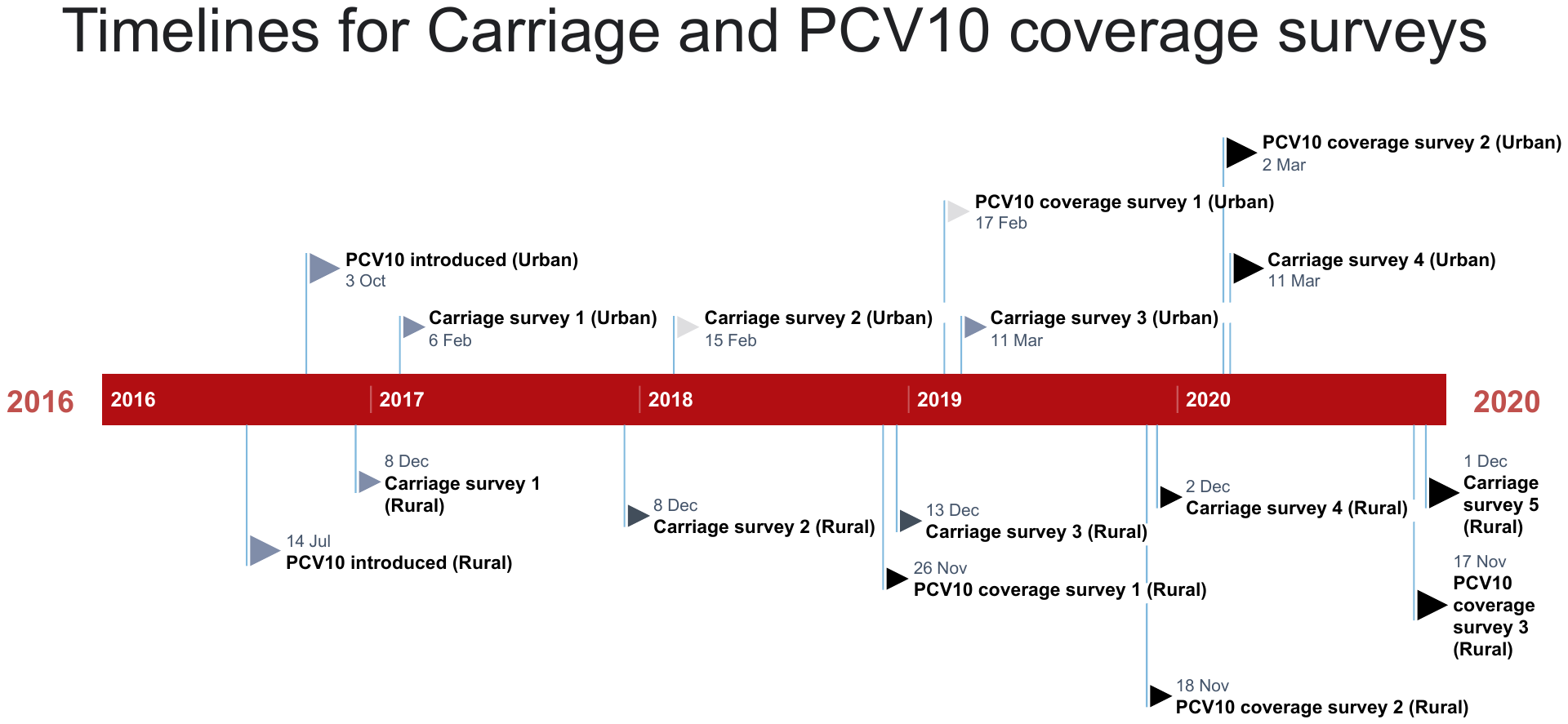

S1 Fig: Timelines for surveys in the two sites
