## Supplementary material for "The impact of introduction of the 10-valent pneumococcal conjugate vaccine (PCV10) on pneumococcal carriage in Nigeria": S1A Table

S1A Table: Numbers of participants invited, consented, and swabbed in each carriage survey in the rural site.

| **Rural** | | | | | | | | | | | | | |
| --- | --- | --- | --- | --- | --- | --- | --- | --- | --- | --- | --- | --- | --- |
|  | Numbers of participants | | | | | | | | | | | | |
|  | 2016* | 2017 | | | 2018 | | | 2019 | | | 2020 | | |
| Age group | swabbed | invited | consented | swabbed | invited | consented | swabbed | invited | consented | swabbed | invited | consented | swabbed |
| <1 | 73 | 100 | 76 | 71 | 130 | 120 | 108 | 100 | 83 | 82 | 100 | 91 | 89 |
| 1-2 | 132 | 120 | 110 | 105 | 110 | 108 | 101 | 160 | 153 | 147 | 150 | 147 | 141 |
| 3-4 | 91 | 110 | 98 | 88 | 110 | 103 | 95 | 150 | 143 | 133 | 120 | 110 | 106 |
| 5-9 | 191 | 130 | 126 | 116 | 110 | 105 | 102 | 140 | 130 | 124 | 140 | 126 | 120 |
| 10-14 | 81 | 130 | 120 | 110 | 150 | 130 | 119 | 120 | 109 | 106 | 160 | 155 | 149 |
| 15-19 | 38 | 120 | 112 | 107 | 110 | 101 | 97 | 110 | 103 | 96 | 100 | 87 | 85 |
| 20-39 | 154 | 110 | 106 | 104 | 130 | 126 | 125 | 110 | 103 | 101 | 130 | 124 | 122 |
| 40-49 | 46 | 100 | 61 | 61 | 110 | 87 | 87 | 80 | 52 | 52 | 80 | 49 | 47 |
| 50-59 | 30 | 60 | 40 | 39 | 100 | 76 | 75 | 80 | 55 | 55 | 60 | 40 | 40 |
| ≥60 | 36 | 100 | 78 | 78 | 110 | 92 | 90 | 100 | 77 | 76 | 100 | 60 | 58 |
| Total | 872 | 1080 | 927 | 879 | 1170 | 1048 | 999 | 1150 | 1008 | 972 | 1140 | 989 | 957 |

* Sampling technique was not age-stratified
