## Supplementary material for "The impact of introduction of the 10-valent pneumococcal conjugate vaccine (PCV10) on pneumococcal carriage in Nigeria": S1B Table

S1B Table: Numbers of participants invited, consented, and swabbed in each carriage survey in the urban site.

| **Urban** | | | | | | | | | | |
| --- | --- | --- | --- | --- | --- | --- | --- | --- | --- | --- |
| Numbers of participants | | | | | | | | | | |
|  | 2017* | 2018 | | | 2019 | | | 2020 | | |
| Age group | swabbed | invited | consented | swabbed | invited | consented | swabbed | invited | consented | swabbed |
| <1 | 109 | 110 | 82 | 76 | 100 | 67 | 65 | 60 | 38 | 37 |
| 1-2 | 141 | 110 | 103 | 100 | 110 | 109 | 105 | 80 | 67 | 67 |
| 3-4 | 85 | 100 | 70 | 68 | 100 | 76 | 73 | 100 | 83 | 81 |
| 5-9 | 161 | 130 | 126 | 119 | 125 | 119 | 117 | 120 | 116 | 116 |
| 10-14 | 75 | 140 | 120 | 117 | 120 | 103 | 102 | 130 | 116 | 115 |
| 15-19 | 20 | 110 | 95 | 92 | 100 | 76 | 76 | 100 | 78 | 78 |
| 20-39 | 130 | 100 | 92 | 91 | 110 | 107 | 106 | 110 | 104 | 104 |
| 40-49 | 57 | 110 | 101 | 100 | 110 | 100 | 99 | 120 | 110 | 110 |
| 50-59 | 66 | 110 | 99 | 98 | 100 | 90 | 90 | 100 | 84 | 84 |
| ≥60 | 75 | 100 | 80 | 80 | 110 | 99 | 99 | 80 | 59 | 59 |
| Total | 919 | 1120 | 968 | 941 | 1085 | 946 | 932 | 1000 | 855 | 851 |

* Sampling technique was not age-stratified
