## Supplementary material for "The impact of introduction of the 10-valent pneumococcal conjugate vaccine (PCV10) on pneumococcal carriage in Nigeria": S2A Fig

Rural

Response (%)

|  | 2017 | 2018 | 2019 | 2020 |
| --- | --- | --- | --- | --- |
| <1 | 76 | 92.3077 | 83 | 91 |
| 1-2 | 91.6667 | 98.1818 | 95.625 | 98 |
| 3-4 | 89.0909 | 93.6364 | 95.3333 | 91.6667 |
| 5-9 | 96.9231 | 95.4545 | 92.8571 | 90 |
| 10-14 | 92.3077 | 86.6667 | 90.8333 | 96.875 |
| 15-19 | 93.3333 | 91.8182 | 93.6364 | 87 |
| 20-39 | 96.3636 | 96.9231 | 93.6364 | 95.3846 |
| 40-49 | 61 | 79.0909 | 65 | 61.25 |
| 50-59 | 66.6667 | 76 | 68.75 | 66.6667 |
| ≥60 | 78 | 83.6364 | 77 | 60 |

S2A Fig: Proportion of invited participants in sampled age groups that consented to be swabbed by survey year in the rural site
