## Supplementary material for "The impact of introduction of the 10-valent pneumococcal conjugate vaccine (PCV10) on pneumococcal carriage in Nigeria": S2B Fig

Urban

Response (%)

|  | 2018 | 2019 | 2020 |
| --- | --- | --- | --- |
| <1 | 74.5455 | 67 | 63.3333 |
| 1-2 | 93.6364 | 99.0909 | 83.75 |
| 3-4 | 70 | 76 | 83 |
| 5-9 | 96.9231 | 95.2 | 96.6667 |
| 10-14 | 85.7143 | 85.8333 | 89.2308 |
| 15-19 | 86.3636 | 76 | 78 |
| 20-39 | 92 | 97.2727 | 94.5455 |
| 40-49 | 91.8182 | 90.9091 | 91.6667 |
| 50-59 | 90 | 90 | 84 |
| ≥60 | 80 | 90 | 73.75 |

S2B Fig: Proportion of invited participants in sampled age groups that consented to be swabbed by survey year in the urban site
