## Supplementary material for "The impact of introduction of the 10-valent pneumococcal conjugate vaccine (PCV10) on pneumococcal carriage in Nigeria": S3B Fig

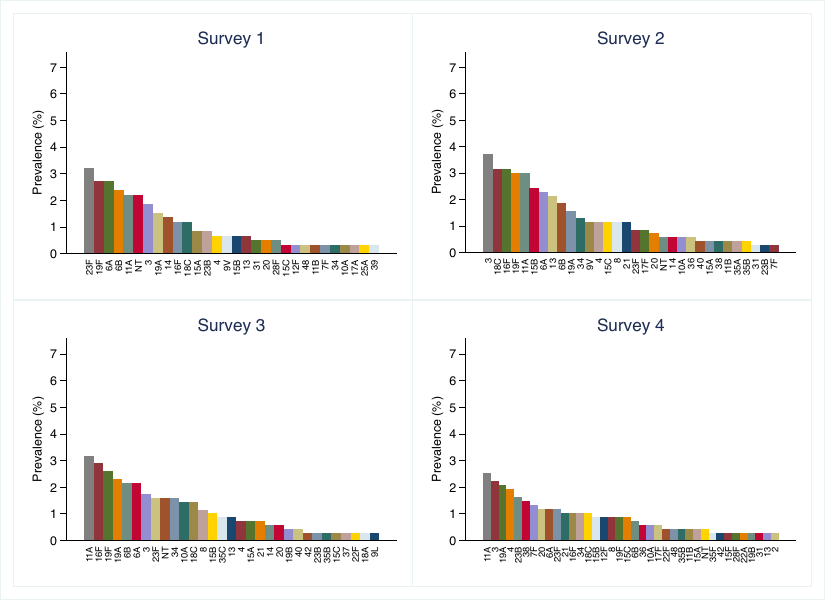

S3B Fig: Serotype-specific carriage prevalence (Serotypes with >1 isolate) among persons aged ≥5 years in the rural site stratified by year of survey
